# Engaging dystonia networks with subthalamic stimulation

**DOI:** 10.1101/2024.05.24.24307896

**Authors:** Konstantin Butenko, Clemens Neudorfer, Till A. Dembek, Barbara Hollunder, Garance M. Meyer, Ningfei Li, Simón Oxenford, Bahne H. Bahners, Bassam Al-Fatly, Roxanne Lofredi, Evan M. Gordon, Nico U.F. Dosenbach, Christos Ganos, Mark Hallett, Philip A. Starr, Jill L. Ostrem, Yiwen Wu, ChenCheng Zhang, Michael D. Fox, Andreas Horn

**Affiliations:** Center for Brain Circuit Therapeutics, Department of Neurology, Brigham & Women’s Hospital, Harvard Medical School, Boston, MA, USA; Department of Neurosurgery, Massachusetts General Hospital, Harvard Medical School, Boston, MA, USA; Department of Neurology, Faculty of Medicine, University of Cologne, Cologne, Germany; Movement Disorders and Neuromodulation Unit, Department of Neurology, Charité – Universitätsmedizin Berlin, Berlin, Germany; Einstein Center for Neurosciences Berlin, Charité – Universitätsmedizin Berlin, Berlin, Germany; Berlin School of Mind and Brain, Humboldt-Universität zu Berlin, Berlin, Germany; Institute of Clinical Neuroscience and Medical Psychology, Medical Faculty and University Hospital Düsseldorf, Heinrich Heine University Düsseldorf, Germany; Department of Neurology, Center for Movement Disorders and Neuromodulation, Medical Faculty and University Hospital Düsseldorf, Heinrich Heine University Düsseldorf, Germany; Mallinckrodt Institute of Radiology, Washington University School of Medicine, St Louis, MO, USA; Department of Neurology, Washington University School of Medicine, St Louis, MO, USA; Department of Biomedical Engineering, Washington University in St. Louis, St Louis, MO, USA; Movement Disorder Clinic, Edmond J. Safra Program in Parkinson’s Disease, Division of Neurology, University of Toronto, Toronto Western Hospital, Toronto, ON, Canada; National Institute of Neurological Disorders and Stroke, National Institutes of Health, Bethesda, MD, USA; Department of Neurological Surgery, University of California, San Francisco, CA, USA; Movement Disorders and Neuromodulation Centre, Department of Neurology, University of California, San Francisco, CA, USA; Department of Neurology & Institute of Neurology, Ruijin Hospital, Shanghai Jiaotong University School of Medicine, Shanghai, China; Department of Neurosurgery, Rujin Hospital, Shanghai Jiaotong University Schools of Medicine, Shanghai, China

## Abstract

Deep brain stimulation is a viable and efficacious treatment option for dystonia. While the internal pallidum serves as the primary target, more recently, stimulation of the subthalamic nucleus (STN) has been investigated. However, optimal targeting within this structure and its complex surroundings have not been studied in depth. Indeed, multiple historical targets that have been used for surgical treatment of dystonia are directly adjacent to the STN. Further, multiple types of dystonia exist, and outcomes are variable, suggesting that not all types would profit maximally from the exact same target. Therefore, a thorough investigation of the neural substrates underlying effects on dystonia symptoms is warranted.

Here, we analyze a multi-center cohort of isolated dystonia patients with subthalamic implantations (*N* = 58) and relate their stimulation sites to improvement of appendicular and cervical symptoms as well as blepharospasm. Stimulation of the ventral oral posterior nucleus of thalamus and surrounding regions was associated with improvement in cervical dystonia, while stimulation of the dorsolateral STN was associated with improvement in limb dystonia and blepharospasm. This dissociation was also evident for structural connectivity, where the cerebellothalamic, corticospinal and pallidosubthalamic tracts were associated with improvement of cervical dystonia, while hyperdirect and subthalamopallidal pathways were associated with alleviation of limb dystonia and blepharospasm. Importantly, a single well-placed electrode may reach the three optimal target sites. On the level of functional networks, improvement of limb dystonia was correlated with connectivity to the corresponding somatotopic regions in primary motor cortex, while alleviation of cervical dystonia was correlated with connectivity to the recently described ‘action-mode’ network that involves supplementary motor and premotor cortex. Our findings suggest that different types of dystonia symptoms are modulated via distinct networks. Namely, appendicular dystonia and blepharospasm are improved with modulation of the basal ganglia, and, in particular, the subthalamic circuitry, including projections from the primary motor cortex. In contrast, cervical dystonia was more responsive when engaging the cerebello-thalamo-cortical circuit, including direct stimulation of ventral thalamic nuclei.

These findings may inform DBS targeting and image-based programming strategies for patient-specific treatment of dystonia.

## Introduction

Building upon decade-long experience of ablative surgeries ^1^, in 1999, the first cases of deep brain stimulation (DBS) in the internal pallidum (GPi) were published ^2,3^. Soon after, the GPi became an established DBS target for various types of dystonia ^4^, including clinical phenotypes with cervical, orofacial and limb manifestations ^5–7^. Yet, the treatment outcome is variable ^8,9^, with the stimulation limited by side-effects such as gait impairment ^10,11^, dysarthria ^12^ and bradykinesia ^13^. More recently, the subthalamic nucleus (STN) has been investigated as an alternative target ^14,15^ with clinical trials showing comparable efficacy ^16,17^. However, while extensive mapping of optimal target sites within the pallidum has been carried out ^18,19^, it remains unclear which exact targets within the STN region provide maximal efficacy. Moreover, the STN occupies a unique anatomical location that is traversed by various thin white matter bundles and resides directly adjacent to multiple gray matter areas. Some of these structures, such as the fields of Forel ^20^ and various nuclei of the thalamus ^21^ have been targeted in electrical stimulation and ablative treatment of dystonia in the past. It could well be that by activating dorsal contacts on electrodes implanted to the STN, one would predominantly modulate other structures, such as the subthalamic area or caudal zona incerta, thalamic nuclei, or the white matter bundles traversing this region, such as ansa and fasciculus lenticulares, comb fibers or cerebellothalamic tract ^22^. We hypothesize that the complex anatomy provides a powerful opportunity for subthalamic stimulation, but also requires precise mapping of this region. Once better understood, it could become possible to engage different circuits affected by dystonia pathophysiology ^23^, potentially in simultaneous fashion, by co-activating different contacts along the same electrode.

Here, we retrospectively investigated optimal targets, tracts and networks based on a comparably large multicenter cohort with heterogeneous dystonic symptoms. We employed previously introduced statistical tools for local mapping as well as structural and functional connectivity analyses ^24^. We focused on three clusters of symptom improvements that were motivated by both clinical experience and literature findings from both historical ^21^ and modern ^19,25,26^ times. Namely, we clustered the available cohort into three partly overlapping subcohorts that presented with predominant baseline symptoms of cervical dystonia, appendicular/limb dystonia (extremities), or blepharospasm (periocular region). By applying the multimodal neuroimaging analysis, we were able to investigate similarities and differences for optimal targeting in STN-DBS for these three types of dystonia on local, tract and network levels.

## Methods

### Patient Cohorts and Imaging

To identify anatomical substrates associated with optimal clinical outcome, we aggregated a total of 78 patients across two independent centers (Shanghai *N* = 65, San Francisco *N* = 13) who underwent STN-DBS for the treatment of isolated therapy-refractory dystonia, which is the largest STN-DBS dystonia dataset studied, to date. Procedures of clinical trials and studies leading to the collection of the data were approved by the institutional review boards at the respective data collection sites in accordance with the Declaration of Helsinki. The STN targeting strategy was not dependent on clinical phenotype. Ages of onset, disease duration and body distribution varied across subjects (*Fig. 1 and Table S4* for inclusion criteria and demographics). Due to the significant difference in *N*, we refrained from comparative subanalyses between cohorts and further addressed the entire dataset. Based on pathophysiological considerations from prior research ^5,19,25,27–31^ and clinical experience, patients were grouped according to their baseline *Burke-Fahn-Marsden Dystonia Rating Scale* (BFMDRS) scores. Due to complex and heterogeneous manifestations of dystonia, we deliberately avoided classifying patients *exclusively*, but assigned them to the groups in overlapping fashion. *Figure 1* graphically illustrates the grouping and summarizes inclusion criteria for each group. We then analyzed clinical improvements of cervical (neck), appendicular (arms and legs) as well as blepharospasm (eyes) items of the BFMDRS after at least three months of DBS ^5^.

**Figure 1:**
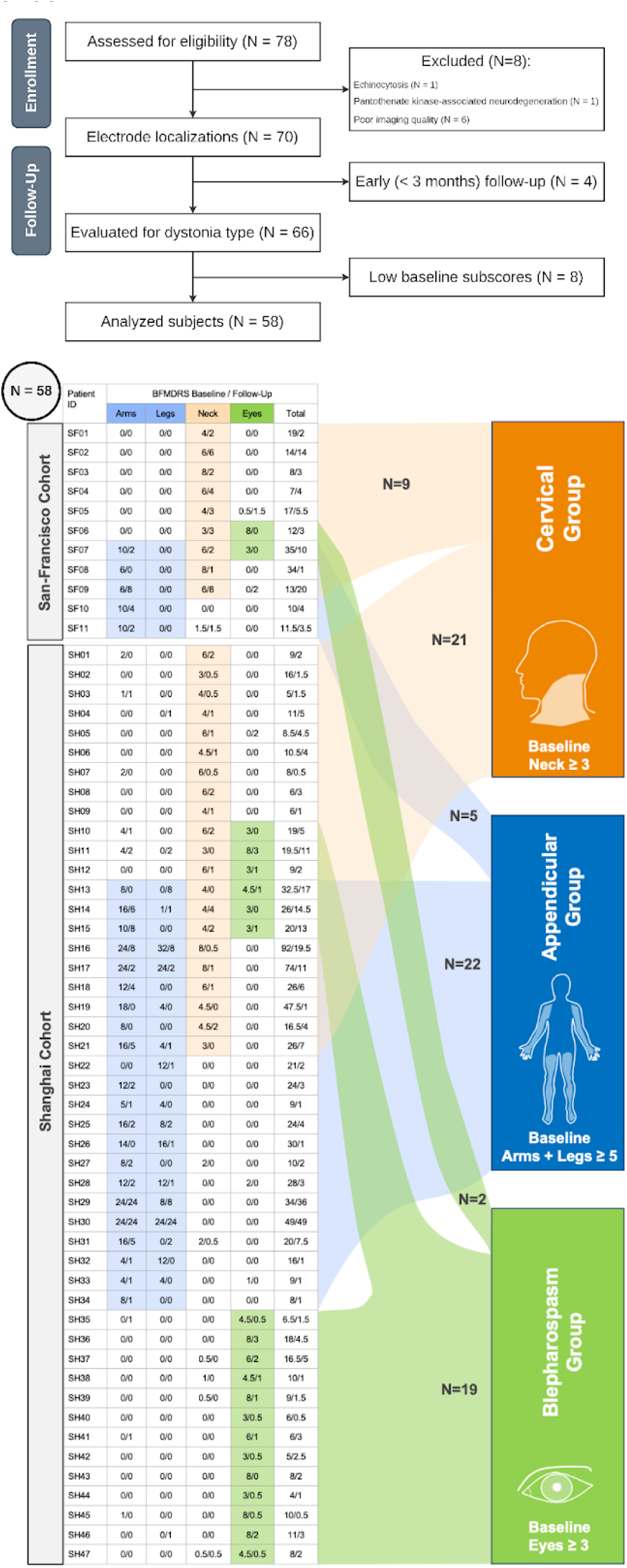
Clinical characteristics and subdivision of patients into three overlapping subcohorts. A: Consort flowchart for inclusion criteria. Patients from two independent cohorts (Shanghai N = 65, San Francisco N = 13) were considered as a single dataset. The final dataset consisted of 58 subjects. B: Classification into dystonic groups based on corresponding baseline subscores. Note that the patients are assigned to groups in non-exclusive fashion.

For every patient, electrode localizations and stimulation volumes were computed using Lead-DBS v3.0 ^24^ as detailed elsewhere ^22^. Briefly, preoperative MRI and postoperative CT/MRI images were first co-registered and then non-linearly warped to ICBM 2009b Nonlinear Asymmetric (‘MNI’) space using Advanced Normalization Tools (ANTs, https://stnava.github.io/ANTs/). Postoperative co-registrations were additionally corrected for brain shift due to possible pneumocephalus ^32^. The non-linear warps were manually refined in the STN region using the *WarpDrive* tool ^33^ in case the automatic delineation of the nucleus was visibly off. Next, the electrode trajectories were reconstructed using either PaCER ^34^ or TRAC/CORE ^35^ algorithms for postoperative CT (*N* = 59) and MRI scans (*N* = 11), respectively (*Figs. 2,3,4, panel A*). Finally, based on clinical stimulation protocols and electrode localizations, distributions of the induced electric fields were computed using the SimBio/FieldTrip pipeline (https://www.mrt.uni-jena.de/simbio/; http://fieldtriptoolbox.org<u>/</u>) ^36^ to solve the static formulation of Maxwell’s equations using the Finite Element Method. The computational domain was discretized into four compartments with distinct electrical conductivities (metal: 10⁸ S/m; insulation: 10⁻¹⁶ S/m; gray matter: 0.33 S/m; white matter: 0.14 S/m) according to the electrode reconstruction and the basal ganglia tissue distribution defined by the DISTAL atlas ^37,38^. The spatial distribution of the electric field magnitude (further referred to as E-field) was subsequently used as the main underlying parameter for the stimulation effect.

**Figure 2:**
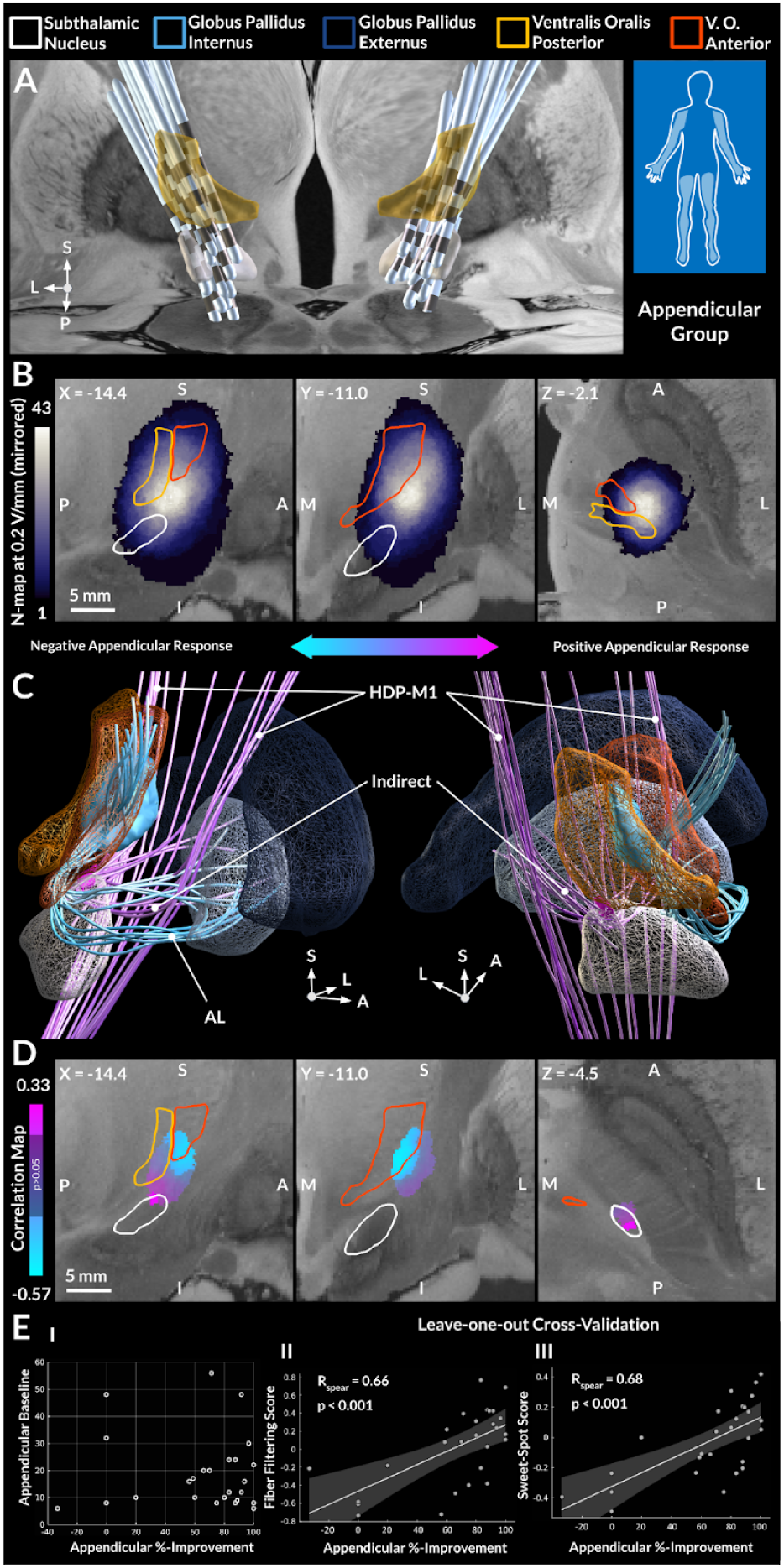
Appendicular Dystonia. A: Electrode reconstructions for patients included in the appendicular group. Note that proximal electrode contacts and stimulation volumes covered the ventral thalamus due to a comparably large contact spacing in the implanted electrodes. B: Distribution of stimulation volumes, defined by electric fields thresholded at the magnitude of 0.2 V/mm. The peak intensity resides in the white matter region dorsolateral to the STN. C: Structural connectivity statistically associated (p<0.05) with appendicular improvement under DBS. Stimulation of the subthalamopallidal (indirect) pathway and the hyperdirect projections from the lower and upper extremity regions of the primary motor cortex (HDP-M1) positively correlated with clinical improvements, while the opposite was observed for the ansa lenticularis (AL). D: Voxel-wise correlation map of the electric field magnitude with stimulation outcome. The sweet-spot, thresholded for significance, localized to the dorsolateral aspect of motor STN, while the sour-spot predominantly resided in the ventral oral anterior nucleus of thalamus. E: Clinical scores and their association with the fiber filtering and sweet-spot scores, quantified by spatial correlations between E-field distributions and sweet-spots / tractograms; Fisher Z-transformation was applied to the sweet-spot scores. To indicate robustness of the association, leave-one-out cross-validation is reported. For 10-fold cross-validations see Fig. S1. Models were also subjected to permutation tests: R = 0.69, p < 0.001 and R = 0.79, p < 0.001 for the sweet-spot and fiber filtering models, respectively.

**Figure 3:**
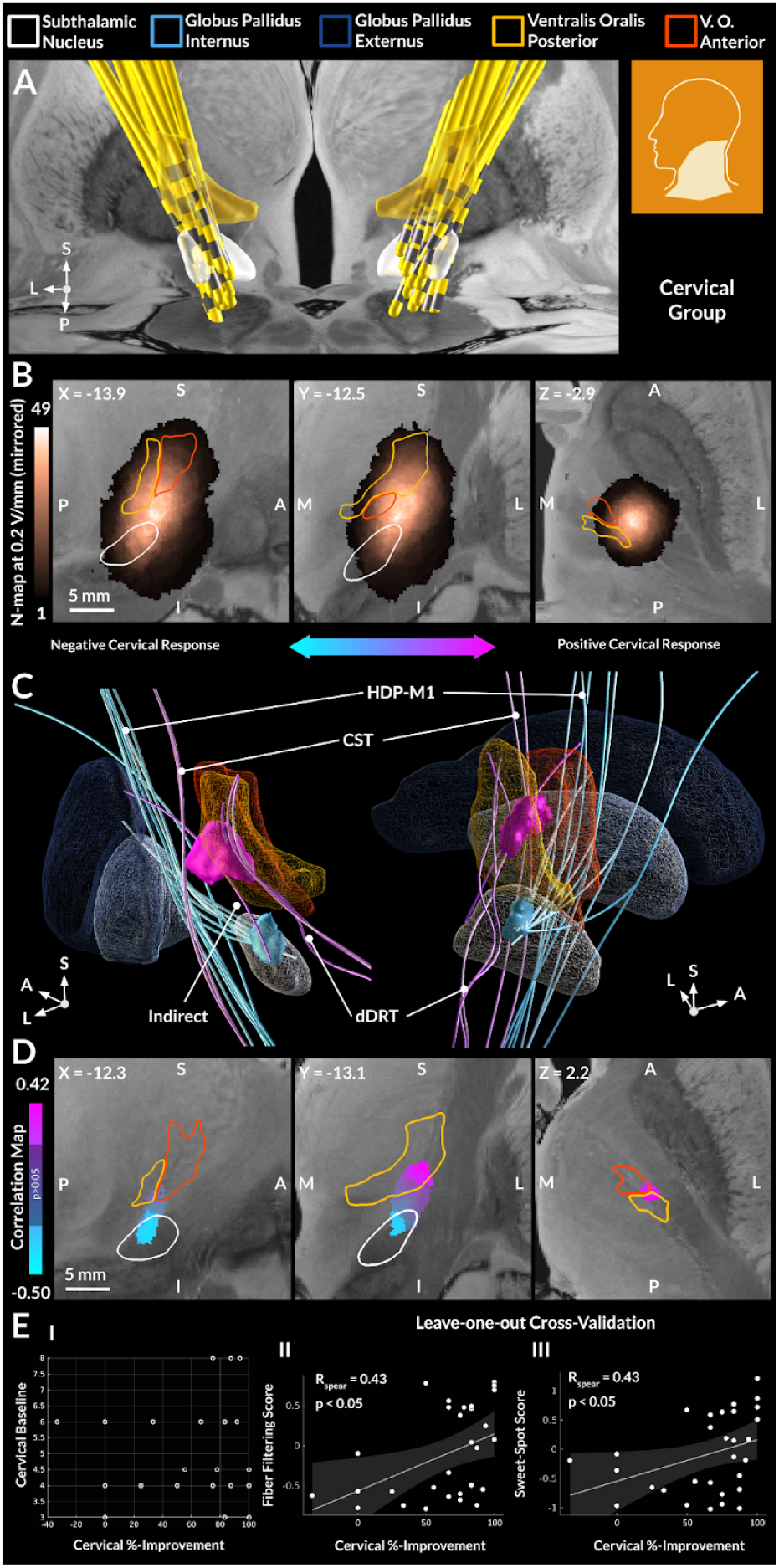
Cervical Dystonia. A: Electrode reconstructions for patients included in the cervical group. Note that proximal electrode contacts and stimulation volumes covered the ventral thalamus due to a comparably large contact spacing in the implanted electrodes. B: Distribution of stimulation volumes, defined by electric fields thresholded at the magnitude of 0.2 V/mm. The peak intensity resides in the white matter region dorsolateral to the STN. C: Structural connectivity statistically associated (p<0.05) with cervical improvement under DBS. Stimulation of the cerebellothalamic (dDRT) and corticospinal tracts (CST), as well as subthalamic afferents from the globus pallidus externus (indirect) positively correlated with clinical improvements, while the opposite was observed for the hyperdirect pathway from the primary motor cortex (HDP-M1). D: Voxel-wise correlation map of the electric field magnitude with stimulation outcome. The sweet-spot, thresholded for significance, primarily localized to the ventral oral posterior nucleus of thalamus, while the sour-spot was identified at the border of the motor and associative STN. E: Clinical scores and their association with the fiber filtering and sweet-spot scores, quantified by spatial correlations between E-field distributions and sweet-spots / tractograms; Fisher Z-transformation was applied to the sweet-spot scores. To indicate robustness of the association, leave-one-out cross-validation is reported. For 10-fold cross-validations see Fig. S1. Models were also subjected to permutation tests: R = 0.50, p = 0.055 and R = 0.62, p < 0.05 for the sweet-spot and fiber filtering models, respectively.

### Analysis of neural correlates

Since DBS affects not only gray, but also white matter and distributed brain networks ^39–41^, optimal DBS targets may best be defined as a triad of i) the specific location associated with optimal outcomes (sweet-spot), ii) the spatial trajectory of white matter tracts associated with optimal outcomes, and iii) the polysynaptic functional brain network associated with optimal outcomes ^42^. Therefore, we analyzed optimal target sites for each type of dystonia on a local, tract- and network level. To do so, we carried out DBS sweet-spot, fiber filtering and network mapping analyses as implemented in Lead-DBS and described in detail elsewhere ^19,22,43–45^. In brief, we conducted a mass-univariate correlation analyses based on the voxel-wise (sweet-spot ^19,45^) and fiber-wise (fiber filtering ^19,22,44^) E-field metric as well as fMRI-based correlation maps (network mapping ^19,44^). For the fiber filtering analysis, relevant streamlines were identified from tracts implemented in the Basal Ganglia Pathway Atlas ^46^. Created by experienced neuroanatomists in a semi-manual fashion, this atlas precisely describes the majority of tracts directly affected by stimulation in the STN region. For network mapping, correlation maps were obtained based on an *N* = 1,000 normative resting state functional connectome ^47,48^, seeded from each E-field to calculate the respective connectivity maps ^49^. Note that all analyses employed Spearman’s correlations due to the non-normal distributions of both E-fields and clinical scores (for the latter, see *Figs. 2,3,4, panel E*). Since clinical improvements were evaluated for bilateral stimulations, we follow previous retrospective studies on dystonia and mirror the stimulation sites between the two hemispheres when defining the models ^18,19^.

**Figure 4:**
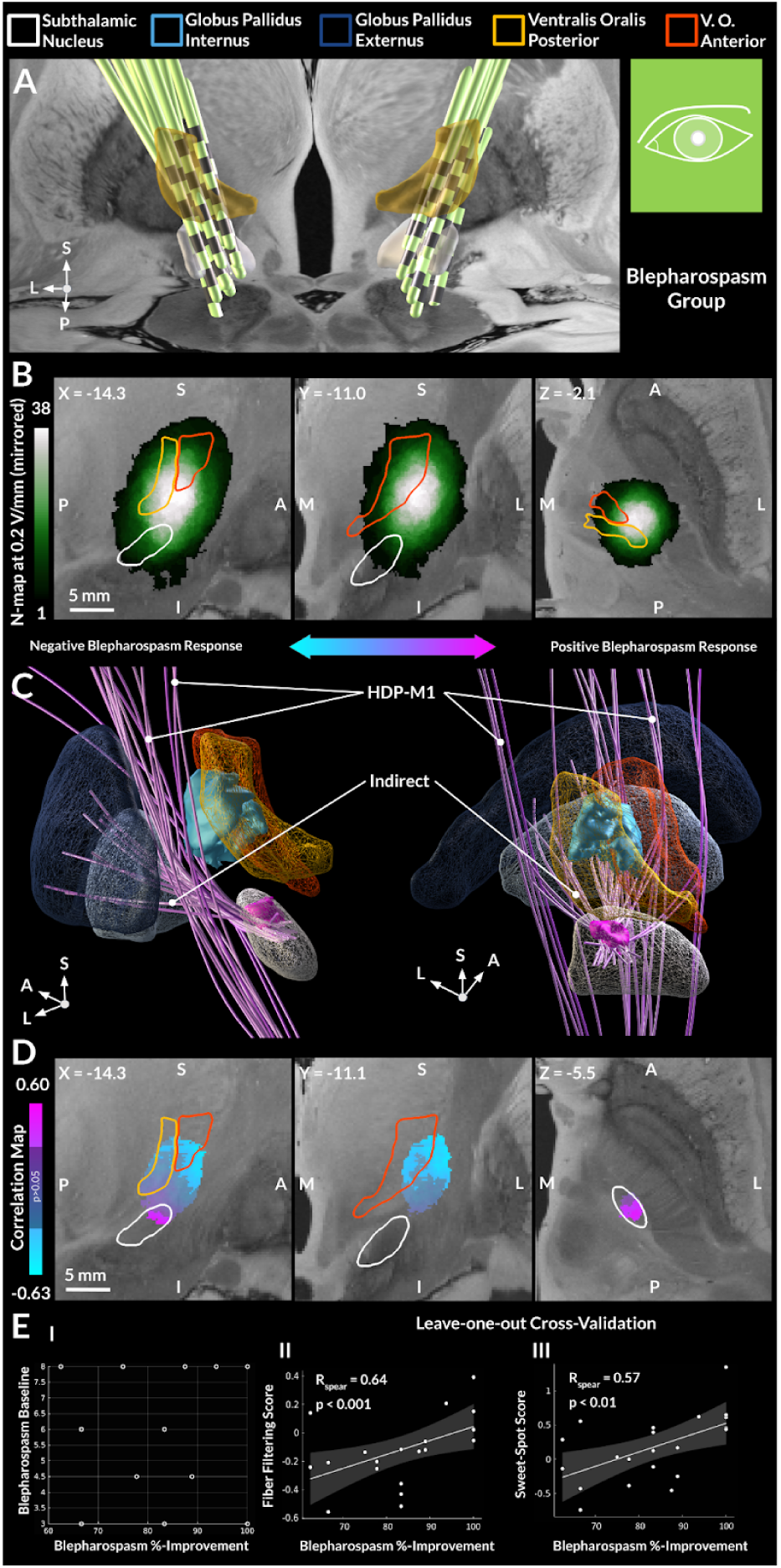
Blepharospasm. A: Electrode reconstructions for patients included in the blepharospasm group. Note that proximal electrode contacts and stimulation volumes covered the ventral thalamus due to a comparably large contact spacing in the implanted electrodes. B: Distribution of stimulation volumes, defined by electric fields thresholded at the magnitude of 0.2 V/mm. The peak intensity resided in the white matter region dorsolateral to the STN. C: Structural connectivity statistically associated (p<0.05) with blepharospasm improvement under DBS. Stimulation of the subthalamopallidal tract (indirect), as well as the hyperdirect pathway from the primary motor cortex (HDP-M1) positively correlated with clinical improvements. D: Voxel-wise correlation map of the electric field magnitude with stimulation outcome. The sweet-spot primarily localized to the motor STN proper, while the sour-spot resided in the capsule and, partially, in the ventral oral nuclei of the thalamus. E: Clinical scores and their association with the fiber filtering and sweet-spot scores, quantified by spatial correlations between E-field distributions and sweet-spots / tractograms; Fisher Z-transformation was applied to the sweet-spot scores. To indicate robustness of the association, leave-one-out cross-validation is reported. For 10-fold cross-validations see Fig. S1. Models were also subjected to permutation tests: R = 0.73, p < 0.001 and R = 0.60, p <0.01 for the sweet-spot and fiber filtering models, respectively.

Such analyses are conducted in a highly multidimensional (voxel or fiber) space prone to overfitting. Therefore, to evaluate the robustness of generated models, we subjected them to leave-one-out (LOO) and 10-fold cross-validations. Here, in each iteration, a part of the data was held-out, and the model was defined using the remaining subjects. Based on this new model, scores for the held-out patients were computed, and the procedure was repeated across all folds / patients. The obtained scores were then correlated with the observed clinical improvement. For the 10-fold cross-validation, we repeated the procedure 10 times (i.e., 100 models were generated in total) to ensure robustness for different configurations of folds. To quantify the possibility of a type I error, we also subjected models to permutation-based testing. For this, a null-distribution was obtained from a set of Spearman’s correlation coefficients calculated after permuting clinical improvements across patients 1,000 times to derive permuted model scores. The null hypothesis was then rejected with a probability estimated by performance of the original model (quantified by the correlation coefficient) against the permuted models (alpha = 0.05). Along the same lines, we evaluated a similarity of sweet-spot maps comparing their correlation coefficient with correlations between the maps of permuted models.

## Results

For demographics and clinical results, see *Fig. 1 and Table S4*. The consort flowchart in *Fig. 1* details exclusions. In brief, out of 78 patients, two subjects were excluded due to a diagnosis of echinocytosis or pantothenate kinase-associated neurodegeneration, respectively. Six subjects were further excluded due to missing or low-quality imaging data. For four subjects, only early (less than three months) follow-ups were available. Eight subjects had low baseline scores in all of the considered groups (see the criteria in *Fig. 1*) and were excluded to avoid bias when analyzing percent improvements. After these exclusions, a total of 58 subjects were retained (29 female, mean age at surgery 40.2 ± 20.1 years, disease duration 6.6 ± 7.6 years, follow-up assessment 1.3 ± 1.2 years after surgery), who were non-exclusively assigned to the appendicular (*N* = 27), cervical (*N* = 30), and blepharospasm (*N* = 21) groups. Four patients qualified for inclusion in all three groups, eight into both cervical and appendicular groups and four patients into cervical and blepharospasm groups. Across subjects, considerable variability in outcomes was observed in the total BFMDRS percent improvement (67.7 ± 28.4%) and body region-specific percent improvement (appendicular 73.7 ± 32.1%, cervical 64.4 ± 34.9%). Notably, the variance in clinical outcome was comparably smaller in the blepharospasm group with patients showing a high response (82.3 ± 13.1%). Neither duration of the disease nor age at surgery significantly correlated with clinical outcomes for any of the considered groups, see *Fig. S5*. Electrodes were localized within the subthalamic region in all patients (*Figs. 2, 3, 4, panel A*). Across the three groups, the distribution of E-fields, binarized at 0.2 V/mm, clearly revealed the highest overlap in the region dorsolateral to the STN, see *Figs. 2,3,4, panel B*. In other words, even though the STN was the surgical target in these patients, chronic stimulation was primarily carried out in the white matter dorsally adjacent to the nucleus.

### Appendicular dystonia

For the appendicular group, stimulation at the dorsolateral aspect of the STN was associated with better outcome, while the “sour-spot”, corresponding to suboptimal improvement, resided predominantly in the ventral oral anterior nucleus (V.o.a.) of thalamus bordering the internal capsule (*Fig. 2, panel D* and *Fig. 5*). These results were robust when subjected to leave-one-out cross-validation (LOO: R = 0.68, p < 0.001, *Fig. 2, panel E-III*), 10-fold cross-validation (see *Fig. S1*), and permutation-based analysis (R = 0.69, p < 0.001). Notably, the “sweet-spot” corresponded to the terminating region of the hyperdirect pathway originating from the primary motor cortex (*Fig. 2, panel C*), whose stimulation was also associated with symptom improvement (LOO: R = 0.66, p < 0.001, *Fig. 2, panel E-II;* perm. test: R = 0.79, p < 0.001; for *10-fold see Fig. S1*). On the other hand, fibers associated with suboptimal improvements belonged to the ansa lenticularis, which accordingly also traversed the “sour-spot” in V.o.a. Finally, on a polysynaptic network level, the functional connectivity analysis demonstrated a correlation of appendicular improvement with stronger connectivity to the precentral gyrus. When repeating this analysis focusing on upper and lower limb items separately, mappings matched the effector sites (arm and leg regions; *Fig. 7, panel B*).

**Figure 5:**
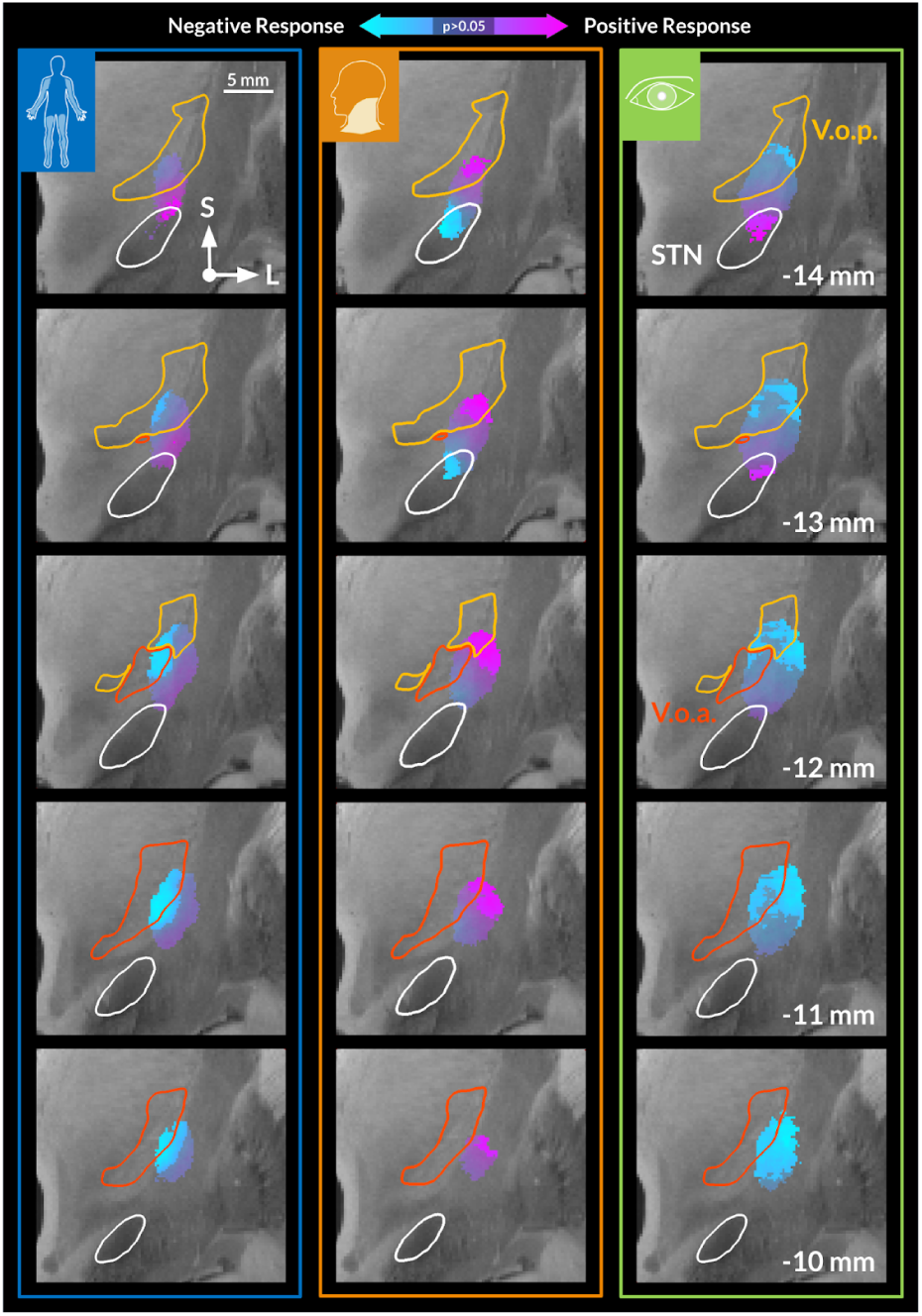
Sweet-spot mappings in the three types of dystonia. The sweet- and sour-spots are computed by correlating distributions of electric fields with clinical outcomes across patients, coronal view. Note a partial inversion of the map for the cervical group, with the optimal stimulation site located in the thalamic and dorsal white matter region. A permutation-based similarity testing indicated a significant positive correlation of the appendicular and blepharospasm maps (R = 0.58, p<0.001), and their non-significant negative correlation with the cervical map (R = -0.09, p = 0.77 and R = -0.37, p = 0.11, respectively).

### Cervical dystonia

A clearly distinct pattern was discovered for improvement of cervical dystonia (*Fig. 3, panel D and Fig. 5*) that primarily mapped to the ventral oral posterior nucleus (V.o.p.) of the thalamus (LOO: R = 0.43, p < 0.05, *Fig. 3, panel E-III*; perm. test: R = 0.50, p = 0.055; for *10-fold see Fig. S1*). Curiously, the sour-spot was identified at the border between the motor and associative subterritories of the STN. While, on a sweet-spot level, this pattern appears as an ‘inversion’ of the results for appendicular dystonia (where the optimal site was in the STN but the suboptimal site in the thalamus), the results were not perfectly inverse; the permutation-based statistics showed that the maps were not significantly more dissimilar than expected by chance (R = -0.09, p = 0.77). In other words, the thalamic sweet-spot for cervical dystonia mapped to a different site than the thalamic sour-spot for appendicular dystonia, and the same was true for the STN (see *Fig. 5* for direct comparison and *Fig. 6* for representative example cases of patients with dystonias involving multiple body regions and their respective improvements). DBS fiber filtering revealed a greater improvement with stimulation of the cerebellothalamic pathway (*Fig. 3, panel C*), as well as the corticospinal tract and a portion of the subthalamic afferents originating from the globus pallidus externus (LOO: R = 0.43, p < 0.05, *Fig. 3, panel E-II;* perm. test: R = 0.62, p < 0.05; for *10-fold see Fig. S1)*. On the level of polysynaptic functional networks, higher improvement was associated with stronger connectivity to the cingulo-opercular ‘action-mode’ network ^50^ (see *Fig. 7),* but not the primary motor cortex, as in the case of appendicular dystonia. A secondary analysis, which repeated the same steps for a more inclusive ‘axial’ improvement (comprising cervical, truncal, and oral improvements), revealed highly similar results, see *Fig. S2*.

**Figure 6:**
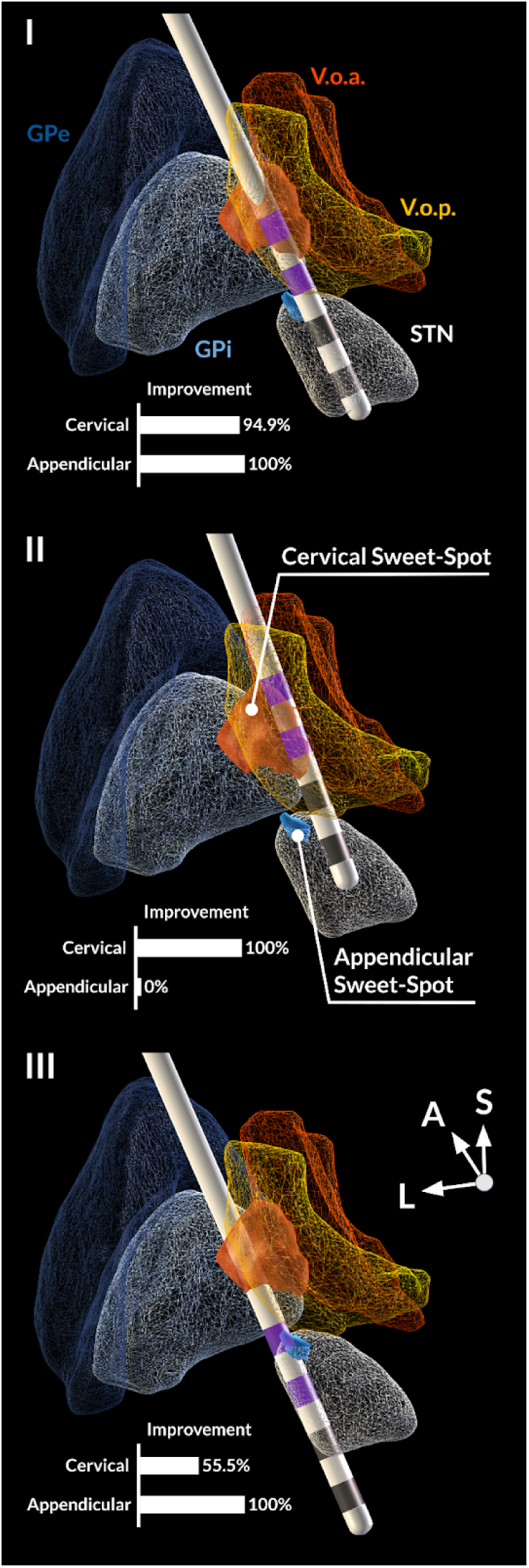
Three representative cases with dystonia involving multiple body regions. Proximity of active contacts (highlighted in purple) to the sweet-spots (blue - appendicular, orange - cervical) reflects the improvement in the corresponding symptoms. The first patient had improvement in both cervical and appendicular symptom clusters. Their two active contacts mapped to both sweet-spots. The second and third patients had strong improvements only in one of the clusters, matching the sweet-spots that were stimulated. Note that the presumably optimal trajectory would coincide with the commonly employed trajectory for DBS in Parkinson’s disease.

**Figure 7:**
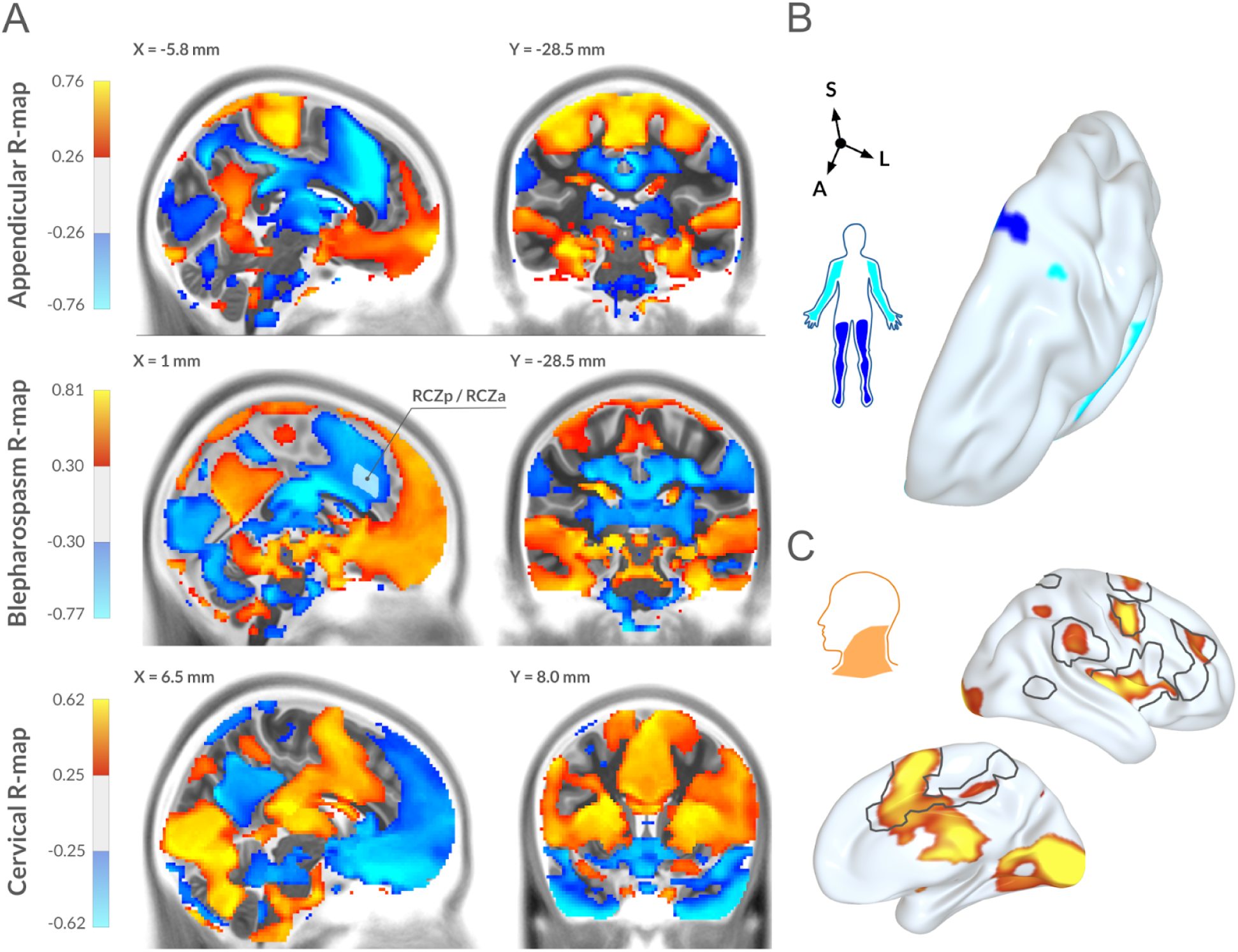
Functional connectivity patterns associated with improvements in the three types of dystonia. A: Maps of symptomatic improvement-associated functional connectivity across groups thresholded at the significance level p < 0.05. B: An analysis of improvement specifically in arms and legs scores reveals a localization of the peaks of the corresponding maps to the somatomotor regions of the primary motor cortex. C: Peaks of the cervical map primarily reside in the cingulo-opercular ‘action-mode’ network ^50^, shown by gray contours.

### Blepharospasm

In contrast to the two previous groups, a high DBS response was observed for blepharospasm treatment in all considered patients, see *Fig. 4, panel E-I.* The overall pattern of structures implied in symptom modulation was similar to the appendicular group (*Fig. 4, panel D a*nd *Fig. 5,* permutation-based similarity R = 0.58, p < 0.001). Namely, sweet-spot mapping identified greater improvement of symptoms in the motor subregion of the STN (LOO: R = 0.57, p < 0.01, *Fig. 4, panel E-III;* perm. test: R = 0.73, p <0.001; for *10-fold see Fig. S1)*, which also matched the termination site for the positively correlated fibers (*Fig. 4, panel C*) of the hyperdirect pathway originating in M1 and the motor subthalamopallidal tract (LOO: R = 0.64, p < 0.001, *Fig. 4, E-II;* perm. test: R = 0.60, p <0.01; for *10-fold see Fig. S1).* Suboptimal results mapped to the internal capsule and partially to the V.o.a. For association of blepharospasm improvement with the sweet-spot and fibers of the appendicular group and vice versa, see *Fig. S6*. On a whole-brain level, the two networks were spatially correlated (R = 0.95), i.e., similar between appendicular dystonia and blepharospasm. Noteworthy to mention that lower connectivity to the rostral posterior and anterior cingulate zones (RCZp / RCZa) was associated with higher alleviation of blepharospasm. These regions had been linked to blinking movements before ^51^.

## Discussion

The following conclusions may be drawn from this study. First, our results show that successful STN-DBS for dystonia may be consistently and robustly mapped to specific sites in the subthalamic nucleus and its surroundings. This was demonstrated by applying mass-univariate statistics and subjecting results to cross-validations and permutation testing. The robustness of our findings supports the general notion that the treatment is not effective by chance. Instead, the outcome depends on engagement of specific neural substrates that cluster to anatomically meaningful regions.

Critically, we demonstrate that most stimulations were programmed in such a way that the STN proper was avoided, and dorsally adjacent structures were predominantly modulated. Given that pallidothalamic projections pass the STN superiorly, and the internal pallidum has been the main DBS target for dystonia, this may seem generally unsurprising. However, our sweet-spot mapping results suggest the STN proper for optimal stimulation in limb dystonia and blepharospasm. At the same time, the ‘sour-spot’ for cervical dystonia also localized to the STN, while stimulating in the V.o.p. nucleus of the thalamus was associated with optimal outcome. On a tract level, in summary, we identified optimal response for appendicular and blepharospasm groups to be associated with basal ganglia projections and its primary motor cortex afferents, while cerebellothalamic fibers seemed to play a larger role in cervical forms of dystonia. Finally, this notion is further underscored by the fMRI analyses, which suggested that better outcomes of DBS for appendicular dystonia and blepharospasm were associated with a stronger functional connectivity to the primary motor cortex. On the other hand, the cervical group predominantly mapped to a recently described ‘action-mode’ network that may be phylogenetically older than the homuncular ‘fine-motor’ networks ^26,50^.

### Rationale for differentiation of the dystonic forms

While our study made an emphasis on affected body regions, the present cohort could have been separated into other groupings, as well. For instance, a common clinical separation is into focal, segmental, and generalized forms of dystonia, or into cervical vs. generalized forms ^18,19^. Furthermore, groupings are often carried out by clinical experts based on physical examinations or video ratings, instead of, as done here, based on BFMDRS items. This being said, most movement disorder specialists will agree that groupings are never perfect, since the disorder can manifest in numerous overlapping ways, and no single established way of grouping for types of dystonia exists. Prior research has shown that different forms of dystonia may respond best to the stimulation of different networks ^19^. Thus, simply analyzing a heterogeneous cohort based on total BFMDRS improvements might not be a viable option and would conflate or even average out improvements in somatotopically organized symptoms. Here, we chose to use an objective grouping based on three specific body regions: limbs (appendicular dystonia), neck (cervical dystonia) and the periocular region (blepharospasm). The following considerations motivated this grouping. First, studies on acquired dystonia reported a differential mapping of lesions in cervical and appendicular dystonias ^25,28,30^. At the same time, blepharospasm might represent a physiologically distinct case of late onset focal dystonia ^52,53^. A parallel perspective on our grouping was motivated by neuroscientific findings: Evidence accumulates that there is a phylogenetically older system that coordinates gross movements versus a more refined motor control system that originated in primates and is associated with fine-motor movements of the limbs ^26,54^. We hypothesized that the former is dominant for axial dystonic symptoms such as postural instability, involuntary abdominal contractions, torsion and oromandibular control impairment, while the latter would be more involved in dystonia of limb movements, such as arms and legs. It seems that the phylogenetically older system (which would make up most of the motor cortex in rodents) was interspersed with so-called “effector” regions more prominently defined in primates, where the primary motor cortex further enhanced direct projections to motor neurons in the spinal cord. In contrast, the remaining parts of primary motor cortex have recently been termed ‘intereffector regions’, functionally forming the newly-described somato-cognitive action network (SCAN) ^26^. Directly adjacent to the SCAN, resides the cingulo-opercular ‘action-mode’ network ^50^, which is functionally associated to the SCAN to a high degree and includes premotor regions. Engagement of the ‘action-mode’ network occurs during increased arousal, processing of instructional cues, action planning, etc ^50^. This two-network approach into ‘effector/pyramidal’ vs. ‘intereffector/extrapyramidal’ motor systems, however, was less clear for blepharospasm, which affects eyes (effectors) primarily manifesting in involuntary eyelid muscle contractions (intereffectors), see ^26^. Therefore, blepharospasm was considered separately.

### Symptom-specific network modulation in dystonia

The identified differential mappings of DBS effects across the three groups of dystonic symptoms may be informative for clinical care: Both local mapping and structural connectivity analyses showed that modulation of the STN circuitry is beneficial for alleviation of appendicular and eye dystonias, while improvement of axial, predominantly cervical, symptoms was observed for more dorsal stimulation (in the thalamic region). To understand this dichotomy, we analyzed the distributed dystonia network. While the basal ganglia have been considered a primary source of the dystonic pathophysiology ^55–57^, there is also evidence that demonstrates involvement of the thalamus, brainstem and cerebellum ^29,57,58^. Crucially, stimulation in the subthalamic area allows a direct engagement of the aforementioned brain regions via structural connectivity. For example, apart from the basal ganglia circuitry, STN-DBS may recruit thalamic afferents, such as pallidothalamic and cerebellothalamic tracts ^59,60^.

The differential mapping of dystonic phenotypes has in part been reported in prior studies. Namely, secondary *limb* dystonia has been linked to incidental basal ganglia lesions ^25,30^. For secondary blepharospasm, a preferential lesion localization was not observed ^25,61^, however, both subthalamic and pallidal stimulations were shown to be effective in the symptom reduction ^62,63^. These findings are now corroborated by the present results showing that alleviation of dystonias occurring in effectors (such as limbs and periocular region) is associated with the subthalamic circuitry accessed via the STN proper, as well as hyperdirect and indirect pathways, (*Figs. 2* and *4*, *panels C and D).* Notably, secondary *hand* dystonia has been observed after thalamic lesions ^25,64–66^, while surgical lesioning of ventral oral nuclei was shown to be effective for treatment of writer’s cramp and musician dystonia ^67–69^.

In contrast, secondary cervical dystonia has been associated with brainstem and cerebellar lesions ^25,28,31^. Management of cervical symptoms has been linked to surgical lesioning of pallidothalamic tracts in Forel’s field H1 ^70^ and stimulation of this region ^20^. Furthermore, optimal stimulation sites dorsal to the STN, including the zona incerta and Forel’s field H2, have been reported in a recent STN-DBS study on cervical dystonia ^71^. In our study, improvement of cervical symptoms mostly mapped to the circuits bypassing the STN. Except for pallidosubthalamic streamlines, the positively associated structural connections represented cerebellothalamic and corticospinal tracts, see *Fig. 3 panel C.* Furthermore, sweet-spot mapping showed the effective stimulation site located dorsal to the STN at the border of V.o.p., Forel’s field H1, zona incerta and the internal capsule. Nearly the same results were obtained when analyzing the axial group, comprising neck, trunk, and mouth dystonias. (*Fig. S2*). Summarizing this evidence, we theorize that the DBS effect on axial and cervical symptoms, in particular, is primarily imposed via circuitry that does not directly involve the STN.

Notably, our results for cervical dystonia emphasize cerebellothalamic and not pallidothalamic projections. The role of the cerebellum, however, is not surprising, when considering dystonia as a disorder that involves sensory components ^23^. Thalamic nuclei that receive kinesthetic cerebellar input, i.e. VIM / V.o.p. ^72^, further relay it to the somatosensory cortex, implicated in dystonic pathophysiology ^31,73^. From this perspective, the therapeutic effect of DBS might be partially explained by modulation of the pathological sensory signal. In turn, a mechanism of action on the sensory rather than the motor domain could explain the slow time course of treatment response, i.e., that effects of DBS often take months to unfold and hence seem to involve effects of plasticity. Furthermore, abnormal cerebellar activation in fMRI was previously reported in patients with cervical dystonia ^74^. It is important, however, to emphasize the anatomical complexity of the region that we identify as the optimal stimulation site for this manifestation. The intricate trajectory of white matter tracts in the fields of Forel and zona incerta might not be comprehensively accounted for in our analysis. For example, in the employed normative pathway atlas ^46^, the pallidothalamic tracts traverse V.o.a. and terminate in VA, as delineated by the DISTAL Atlas ^37,38^. On the other hand, pallidal projections were also reported for the V.o.p. / VLa region ^75^, which overlaps with the optimal stimulation site discovered in our study. Hence, an exclusive association of the therapeutic effect with either cerebellothalamic or pallidothalamic projections remains disputable. Nevertheless, the present results as well as previous studies emphasize the role of the white matter region dorsal to the STN for alleviation of axial and cervical symptoms.

Our results may be helpful to guide clinical practice. At first glance, seemingly inverted optimal stimulation sites for appendicular dystonia / blepharospasm and cervical dystonia might imply a limited performance of DBS for generalized manifestations. However, this is generally not the case ^9,16^ (though see ^76^), and in our dataset multiple subjects assigned to both cervical and appendicular groups demonstrated a good response in both components (*N* = 8 / 12 above 50% improvement, also see *Fig. 6* for the representative cases). This fact reiterates the notion that “millimeters matter” in DBS, with more beneficial outcomes observed for leads properly placed in motor STN and upper contacts reaching V.o.p. Luckily, such a trajectory coincides with the surgical practice when targeting STN in patients with Parkinson’s disease, so a high accuracy is expected from experienced DBS centers. Furthermore, employment of directional leads with an extended contact array is advisable, since precise surgical targeting of V.o.p. is challenging. The presented results also have an important implication for imaging-based programming, which is especially relevant in DBS for dystonia, since treatment response is often not immediate. According to the local and structural mapping of symptom improvements, patients with predominantly cervical / axial dystonia would have better outcomes when stimulated at the proximal contacts (the V.o.p. and adjacent white matter), while generalized and periocular manifestations would respond better to activating distal contacts (within the STN). During STN-DBS programming, one should consider possible provocations of affective symptoms ^77^ and dyskinesias ^63,78^. The latter might be resolved by a slower increase in the stimulation amplitude and selection of dorsal contacts ^78,79^.

## Limitations

The following limitations should be recognized when interpreting our results. First, the study has been conducted on a retrospective dataset composed of two cohorts, with a total of 58 subjects and three groups of dystonia subtypes ranging from 21 to 30 subjects. Importantly, results could not be validated on an additional held-out test set due to the scarce availability of dystonia datasets with STN implantations. Independent retrospective validation or, ideally, prospective trials would be required to confirm our findings. At the same time, robust cross-validation results indicate a potential for the findings to generalize to unseen data.

Second, severity of symptoms and their localization are interpreted in the context of BFMDRS scores, available for both cohorts, while Toronto Western Spasmodic Torticollis Rating Scale (not available for most of the subjects) could provide additional detail on the cervical component. The BFMDRS scale also does not explicitly evaluate hand dystonia, so this subtype was not investigated. Speech and swallowing components could not be comprehensively addressed due to the low number of patients experiencing this symptom at baseline (*N* = 12 with baseline ≥ 3 points).

Third, the acquired patient imaging was of heterogeneous modality and quality. This inherently affects electrode localization accuracy, despite our advanced image processing that includes multispectral non-linear warping, manual refinement ^33^, brain shift correction ^32^ and phantom-validated trajectory reconstruction ^34^. This limitation also applies to our group-level analysis of electric fields conducted in the common, i.e., MNI space based on non-linear warps of patient scans. Furthermore, for structural and functional connectivity analyses, normative connectomes were employed that do not reflect pathological brain alterations in dystonic patients. Despite using a highly refined anatomical pathway atlas ^46^, some tracts or their collaterals might be missing, obstructing unequivocal interpretation of the results.

Finally, DBS effect on the neural tissue is quantified here by continuous values of extracellular electric field magnitude. We deliberately opted for this metric to incorporate the probabilistic impact of the stimulation in our mass-univariate rank-correlation analyses. For more accurate elaboration on white matter recruitment, pathway activation modeling can be employed ^80^, which, however, requires various assumptions on the axonal morphology and volume conductor complexity.

## Conclusion

The potential of subthalamic stimulation for treatment of dystonia has been previously demonstrated ^15,16,63^, and our study now provides neuroimaging evidence underlying improvement and consistency of optimal stimulation sites. We discover a differential mapping of neural substrates mediating stimulation effects on dystonic symptoms that occur in limbs and eyes vs. cervical / axial presentation. For the former symptoms, effective stimulation directly engaged the subthalamic circuitry, including hyperdirect and indirect pathways. Furthermore, better response for limb dystonia was associated with higher functional connectivity to the primary motor cortex with a somatotopic differentiation. At the same time, improvements of cervical symptoms were associated with more dorsal stimulation of thalamic nuclei and passing fibers, including the cerebellothalamic pathway, with the functional connectivity to the cingulo-opercular ‘action-mode’ network. Importantly, a well-placed STN-DBS electrode may engage both of these substrates simultaneously, potentially facilitating alleviation of distributed symptoms. Our results provide a starting point for more deliberate targeting and programming of subthalamic stimulation for different types of dystonia.

## Data Availability

The code necessary to reproduce the findings of the study is available at https://github.com/Kinway25/leaddbs/tree/STN_DBS_DYT. Upon acceptance, the data necessary to reproduce the findings will be deposited in a separate repository following requirements of the publisher. The raw data are not publicly available due to patient privacy restrictions, but can be requested from the authors within the scope of a data sharing agreement.

## References

1. Gross, R. E. What Happened to Posteroventral Pallidotomy for Parkinson’s Disease and Dystonia? Neurotherapeutics 5, 281–293 (2008).

2. Coubes, P. et al. Treatment of early-onset generalized dystonia by chronic bilateral stimulation of the internal globus pallidus. Apropos of a case. Neurochirurgie. 45, 139–144 (1999).

3. Krauss, J. K., Pohle, T., Weber, S., Ozdoba, C. & Burgunder, J. M. Bilateral stimulation of globus pallidus internus for treatment of cervical dystonia. Lancet Lond. Engl. 354, 837–838 (1999).

4. Kupsch, A. et al. Pallidal Deep-Brain Stimulation in Primary Generalized or Segmental Dystonia. N. Engl. J. Med. 355, 1978–1990 (2006).

5. Volkmann, J. et al. Pallidal neurostimulation in patients with medication-refractory cervical dystonia: a randomised, sham-controlled trial. Lancet Neurol. 13, 875–884 (2014).

6. Reese, R. & Volkmann, J. Deep Brain Stimulation for the Dystonias: Evidence, Knowledge Gaps, and Practical Considerations. Mov. Disord. Clin. Pract. 4, 486–494 (2017).

7. Hao, Q. et al. Pallidal deep brain stimulation in primary Meige syndrome: clinical outcomes and psychiatric features. J. Neurol. Neurosurg. Psychiatry 91, 1343–1348 (2020).

8. Volkmann, J. et al. Pallidal deep brain stimulation in patients with primary generalised or segmental dystonia: 5-year follow-up of a randomised trial. Lancet Neurol. 11, 1029–1038 (2012).

9. Holloway, K. L. et al. Deep brain stimulation for dystonia: a meta-analysis. Neuromodulation J. Int. Neuromodulation Soc. 9, 253–261 (2006).

10. Mahlknecht, P. et al. Parkinsonian signs in patients with cervical dystonia treated with pallidal deep brain stimulation. Brain J. Neurol. 141, 3023–3034 (2018).

11. Schrader, C. et al. GPi-DBS may induce a hypokinetic gait disorder with freezing of gait in patients with dystonia. Neurology 77, 483–488 (2011).

12. Meoni, S. et al. Pallidal deep brain stimulation for dystonia: a long term study. J. Neurol. Neurosurg. Psychiatry 88, 960–967 (2017).

13. Berman, B. D., Starr, P. A., Marks, W. J. & Ostrem, J. L. Induction of Bradykinesia with Pallidal Deep Brain Stimulation in Patients with Cranial-Cervical Dystonia. Stereotact. Funct. Neurosurg. 87, 37–44 (2009).

14. Sun, B., Chen, S., Zhan, S., Le, W. & Krahl, S. E. Subthalamic nucleus stimulation for primary dystonia and tardive dystonia. in *Operative Neuromodulation: Volume 2: Neural Networks Surgery* (eds. Sakas, D. E. & Simpson, B. A.) 207–214 (Springer, Vienna, 2007). doi:10.1007/978-3-211-33081-4_23.

15. Ostrem, J. L. et al. Subthalamic nucleus deep brain stimulation in primary cervical dystonia. Neurology 76, 870–878 (2011).

16. Schjerling, L. et al. A randomized double-blind crossover trial comparing subthalamic and pallidal deep brain stimulation for dystonia. J. Neurosurg. 119, 1537–1545 (2013).

17. Hock, A. N. et al. A randomised double-blind controlled study of Deep Brain Stimulation for dystonia in STN or GPi - A long term follow-up after up to 15 years. Parkinsonism Relat. Disord. 96, 74–79 (2022).

18. Reich, M. M. et al. Probabilistic mapping of the antidystonic effect of pallidal neurostimulation: a multicentre imaging study. Brain J. Neurol. 142, 1386–1398 (2019).

19. Horn, A. et al. Optimal deep brain stimulation sites and networks for cervical vs. generalized dystonia. Proc. Natl. Acad. Sci. U. S. A. 119, e2114985119 (2022).

20. Mundinger, F. [New stereotactic treatment of spasmodic torticollis with a brain stimulation system (author’s transl)]. Med. Klin. 72, 1982–1986 (1977).

21. Hassler, R., Riechert, T., Mundinger, F., Umbach, W. & Ganglberger, J. A. Physiological observations in stereotaxic operations in extrapyramidal motor disturbances. Brain J. Neurol. 83, 337–350 (1960).

22. Hollunder, B. et al. Mapping Dysfunctional Circuits in the Frontal Cortex Using Deep Brain Stimulation. 2023.03.07.23286766 Preprint at 10.1101/2023.03.07.23286766 (2023).

23. Kaji, R., Bhatia, K. & Graybiel, A. M. Pathogenesis of dystonia: is it of cerebellar or basal ganglia origin? J. Neurol. Neurosurg. Psychiatry 89, 488–492 (2018).

24. Neudorfer, C. et al. Lead-DBS v3.0: Mapping deep brain stimulation effects to local anatomy and global networks. NeuroImage 268, 119862 (2023).

25. Corp, D. T. et al. Clinical and Structural Findings in Patients With Lesion-Induced Dystonia: Descriptive and Quantitative Analysis of Published Cases. Neurology 99, e1957–e1967 (2022).

26. Gordon, E. M. et al. A somato-cognitive action network alternates with effector regions in motor cortex. Nature 617, 351–359 (2023).

27. Lee, M. S. & Marsden, C. D. Movement disorders following lesions of the thalamus or subthalamic region. Mov. Disord. Off. J. Mov. Disord. Soc. 9, 493–507 (1994).

28. LeDoux, M. S. & Brady, K. A. Secondary cervical dystonia associated with structural lesions of the central nervous system. Mov. Disord. Off. J. Mov. Disord. Soc. 18, 60–69 (2003).

29. Neychev, V. K., Gross, R. E., Lehéricy, S., Hess, E. J. & Jinnah, H. A. The functional neuroanatomy of dystonia. Neurobiol. Dis. 42, 185–201 (2011).

30. Liuzzi, D., Gigante, A. F., Leo, A. & Defazio, G. The anatomical basis of upper limb dystonia: lesson from secondary cases. Neurol. Sci. Off. J. Ital. Neurol. Soc. Ital. Soc. Clin. Neurophysiol. 37, 1393–1398 (2016).

31. Corp, D. T. et al. Network localization of cervical dystonia based on causal brain lesions. Brain J. Neurol. 142, 1660–1674 (2019).

32. Horn, A. et al. Lead-DBS v2: Towards a comprehensive pipeline for deep brain stimulation imaging. NeuroImage 184, 293–316 (2019).

33. Oxenford, S. et al. WarpDrive: Improving spatial normalization using manual refinements. Med. Image Anal. 91, 103041 (2024).

34. Husch, A., V. Petersen, M., Gemmar, P., Goncalves, J. & Hertel, F. PaCER - A fully automated method for electrode trajectory and contact reconstruction in deep brain stimulation. NeuroImage Clin. 17, 80–89 (2018).

35. Horn, A. & Kühn, A. A. Lead-DBS: A toolbox for deep brain stimulation electrode localizations and visualizations. NeuroImage 107, 127–135 (2015).

36. Vorwerk, J., Oostenveld, R., Piastra, M. C., Magyari, L. & Wolters, C. H. The FieldTrip-SimBio pipeline for EEG forward solutions. Biomed. Eng. OnLine 17, 37 (2018).

37. Chakravarty, M. M., Bertrand, G., Hodge, C. P., Sadikot, A. F. & Collins, D. L. The creation of a brain atlas for image guided neurosurgery using serial histological data. NeuroImage 30, 359–376 (2006).

38. Ewert, S. et al. Toward defining deep brain stimulation targets in MNI space: A subcortical atlas based on multimodal MRI, histology and structural connectivity. NeuroImage 170, 271–282 (2018).

39. Ranck, J. B. Which elements are excited in electrical stimulation of mammalian central nervous system: a review. Brain Res. 98, 417–440 (1975).

40. Nowak, L. G. & Bullier, J. Axons, but not cell bodies, are activated by electrical stimulation in cortical gray matterII. Evidence from selective inactivation of cell bodies and axon initial segments. Exp. Brain Res. 118, 489–500 (1998).

41. Knight, E. J. et al. Motor and Nonmotor Circuitry Activation Induced by Subthalamic Nucleus Deep Brain Stimulation in Patients With Parkinson Disease: Intraoperative Functional Magnetic Resonance Imaging for Deep Brain Stimulation. Mayo Clin. Proc. 90, 773–785 (2015).

42. Horn, A. & Fox, M. D. Opportunities of connectomic neuromodulation. NeuroImage 221, 117180 (2020).

43. Li, N. et al. A unified connectomic target for deep brain stimulation in obsessive-compulsive disorder. Nat. Commun. 11, 3364 (2020).

44. Ríos, A. S. et al. Optimal deep brain stimulation sites and networks for stimulation of the fornix in Alzheimer’s disease. Nat. Commun. 13, 7707 (2022).

45. Meyer, G. M. et al. Deep Brain Stimulation for Obsessive-Compulsive Disorder: Optimal Stimulation Sites. Biol. Psychiatry (2023) doi:10.1016/j.biopsych.2023.12.010.

46. Petersen, M. V. et al. Holographic Reconstruction of Axonal Pathways in the Human Brain. Neuron 104, 1056–1064.e3 (2019).

47. Thomas Yeo, B. T., et al. The organization of the human cerebral cortex estimated by intrinsic functional connectivity. J. Neurophysiol. 106, 1125–1165 (2011).

48. Holmes, A. J. et al. Brain Genomics Superstruct Project initial data release with structural, functional, and behavioral measures. Sci. Data 2, 150031 (2015).

49. Horn, A. et al. Connectivity Predicts deep brain stimulation outcome in P arkinson disease. Ann. Neurol. 82, 67–78 (2017).

50. Dosenbach, N. U. F., Raichle, M. & Gordon, E. M. The brain’s cingulo-opercular action-mode network. (2024) doi:10.31234/osf.io/2vt79.

51. Hanakawa, T., Dimyan, M. A. & Hallett, M. The representation of blinking movement in cingulate motor areas: a functional magnetic resonance imaging study. Cereb. Cortex N. Y. N 1991 18, 930–937 (2008).

52. O’Riordan, S. et al. Age at onset as a factor in determining the phenotype of primary torsion dystonia. Neurology 63, 1423–1426 (2004).

53. Quartarone, A. & Ruge, D. How Many Types of Dystonia? Pathophysiological Considerations. Front. Neurol. 9, 12 (2018).

54. Woolsey, C. N. et al. Patterns of localization in precentral and ‘supplementary’ motor areas and their relation to the concept of a premotor area. Res. Publ. - Assoc. Res. Nerv. Ment. Dis. 30, 238–264 (1952).

55. Galardi, G. et al. Basal ganglia and thalamo-cortical hypermetabolism in patients with spasmodic torticollis. Acta Neurol. Scand. 94, 172–176 (1996).

56. Naumann, M. et al. Imaging the pre- and postsynaptic side of striatal dopaminergic synapses in idiopathic cervical dystonia: a SPECT study using [123I] epidepride and [123I] beta-CIT. Mov. Disord. Off. J. Mov. Disord. Soc. 13, 319–323 (1998).

57. Lehéricy, S., Tijssen, M. A. J., Vidailhet, M., Kaji, R. & Meunier, S. The anatomical basis of dystonia: current view using neuroimaging. Mov. Disord. Off. J. Mov. Disord. Soc. 28, 944–957 (2013).

58. Batla, A. et al. The role of cerebellum in patients with late onset cervical/segmental dystonia?--evidence from the clinic. Parkinsonism Relat. Disord. 21, 1317–1322 (2015).

59. Neudorfer, C. & Maarouf, M. Neuroanatomical background and functional considerations for stereotactic interventions in the H fields of Forel. Brain Struct. Funct. 223, 17–30 (2018).

60. Butenko, K. et al. Linking profiles of pathway activation with clinical motor improvements – A retrospective computational study. NeuroImage Clin. 36, 103185 (2022).

61. Khooshnoodi, M. A., Factor, S. A. & Jinnah, H. A. Secondary Blepharospasm Associated with Structural Lesions of the Brain. J. Neurol. Sci. 331, 98–101 (2013).

62. Jeong, S., Huh, R. & Jang, I. Outcomes of pallidal deep brain stimulation for treating pure blepharospasm. J. Korean Soc. Stereotact. Funct. Neurosurg. 18, 72–80 (2022).

63. Kilbane, C. & Ostrem, J. L. Subthalamic Nucleus Deep Brain Stimulation for Dystonia: Evidence, Pros and Cons. Dystonia 1, 10609 (2022).

64. Huang, G. S., Chen, H. Y. & Tsau, J. C. Thalamic hand: a late onset sequela of stroke and its influence on physical function after rehabilitation: two cases report. Gaoxiong Yi Xue Ke Xue Za Zhi 8, 108–112 (1992).

65. Ghika, J., Bogousslavsky, J., Henderson, J., Maeder, P. & Regli, F. The ‘jerky dystonic unsteady hand’: a delayed motor syndrome in posterior thalamic infarctions. J. Neurol. 241, 537–542 (1994).

66. Deleu, D., Lagopoulos, M. & Louon, A. Thalamic hand dystonia: an MRI anatomoclinical study. Acta Neurol. Belg. 100, 237–241 (2000).

67. Taira, T. & Hori, T. Stereotactic ventrooralis thalamotomy for task-specific focal hand dystonia (writer’s cramp). Stereotact. Funct. Neurosurg. 80, 88–91 (2003).

68. Horisawa, S. et al. Safety and long-term efficacy of ventro-oral thalamotomy for focal hand dystonia. Neurology 92, e371–e377 (2019).

69. Horisawa, S. et al. Magnetic Resonance-Guided Focused Ultrasound Thalamotomy for Focal Hand Dystonia: A Pilot Study. Mov. Disord. Off. J. Mov. Disord. Soc. 36, 1955–1959 (2021).

70. Horisawa, S. et al. Unilateral pallidothalamic tractotomy at Forel’s field H1 for cervical dystonia. Ann. Clin. Transl. Neurol. 9, 478–487 (2022).

71. Yin, F. et al. Bilateral subthalamic nucleus deep brain stimulation for refractory isolated cervical dystonia. Sci. Rep. 12, 7678 (2022).

72. King, N. K. K. et al. Microelectrode recording findings within the tractography-defined ventral intermediate nucleus. J. Neurosurg. 126, 1669–1675 (2017).

73. Inoue, K. et al. Disinhibition of the somatosensory cortex in cervical dystonia-decreased amplitudes of high-frequency oscillations. Clin. Neurophysiol. Off. J. Int. Fed. Clin. Neurophysiol. 115, 1624–1630 (2004).

74. Prudente, C. N. et al. A Functional Magnetic Resonance Imaging Study of Head Movements in Cervical Dystonia. Front. Neurol. 7, (2016).

75. Gallay, M. N., Jeanmonod, D., Liu, J. & Morel, A. Human pallidothalamic and cerebellothalamic tracts: anatomical basis for functional stereotactic neurosurgery. Brain Struct. Funct. 212, 443–463 (2008).

76. Fan, H., Zheng, Z., Yin, Z., Zhang, J. & Lu, G. Deep Brain Stimulation Treating Dystonia: A Systematic Review of Targets, Body Distributions and Etiology Classifications. Front. Hum. Neurosci. 15, 757579 (2021).

77. Volkmann, J., Daniels, C. & Witt, K. Neuropsychiatric effects of subthalamic neurostimulation in Parkinson disease. Nat. Rev. Neurol. 6, 487–498 (2010).

78. Wang, N. et al. Stimulation-Induced Dyskinesia After Subthalamic Nucleus Deep Brain Stimulation in Patients With Meige Syndrome. Neuromodulation J. Int. Neuromodulation Soc. 24, 286–292 (2021).

79. Bledsoe, I. O. et al. Phenomenology and Management of Subthalamic Stimulation-Induced Dyskinesia in Patients With Isolated Dystonia. Mov. Disord. Clin. Pract. 7, 548–551 (2020).

80. Gunalan, K. et al. Creating and parameterizing patient-specific deep brain stimulation pathway-activation models using the hyperdirect pathway as an example. PloS One 12, e0176132 (2017).

