## Supplementary Material for "Engaging dystonia networks with subthalamic stimulation"

### Supplementary Figure 1: Boxplots for 10 shuffles of 10-fold cross-validations

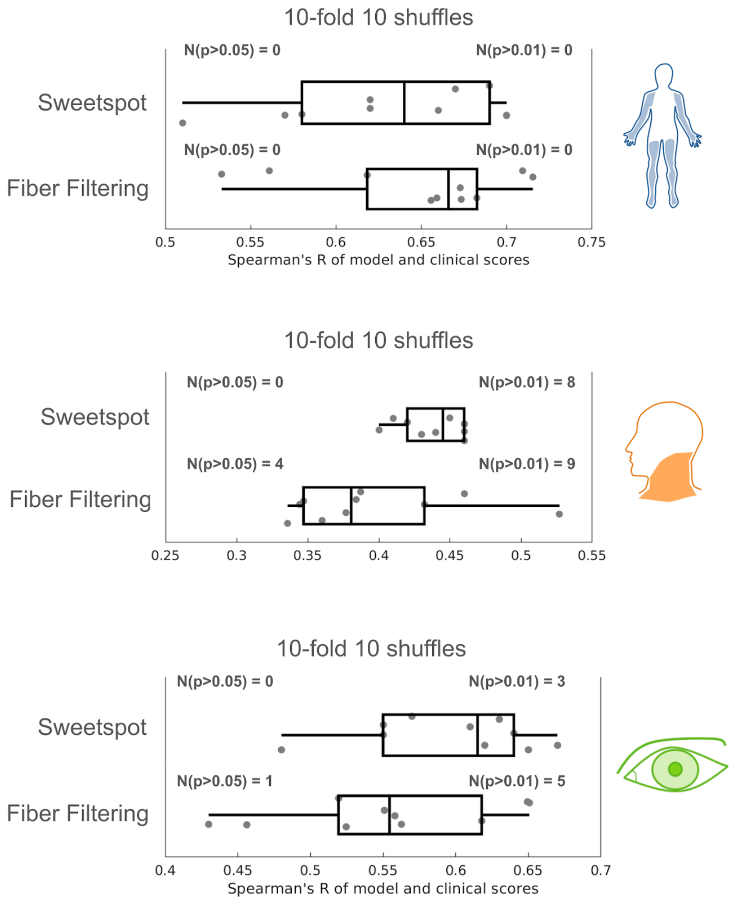

### Supplementary Figure 2: Neural substrate correlated with axial improvement (*N* = 26)

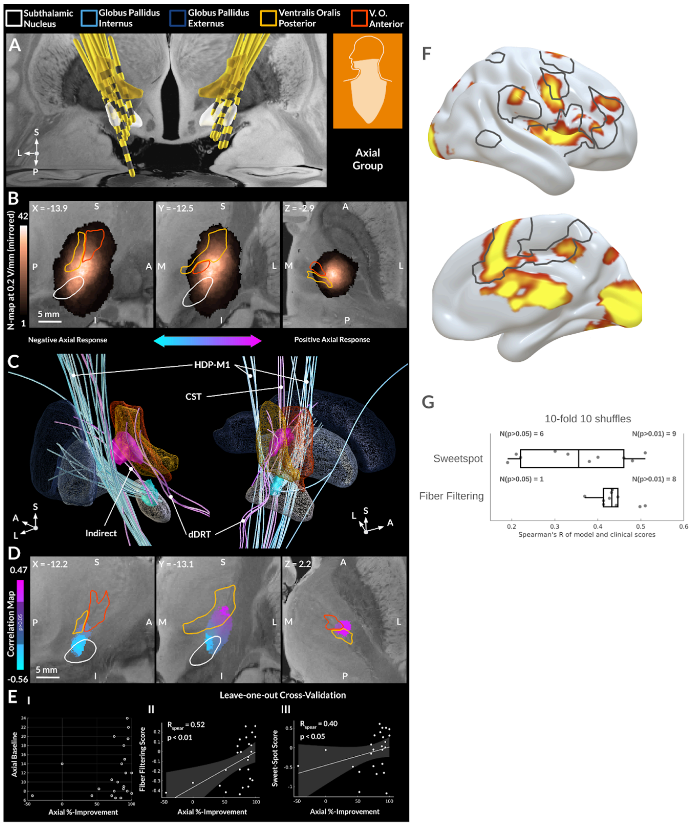


***Axial Dystonia:*** *A total of 26 patients, who had > 6 points in the BFMDRS neck, trunk and mouth items, were included in the axial group. A: Electrode reconstructions for patients included in the axial group. Note that proximal electrode contacts and stimulation volumes covered the ventral thalamus. B: Distribution of patients' stimulation volumes, defined by electric fields thresholded at the magnitude of 0.2 V/mm. The peak intensity resides in the white matter region dorsolateral to the STN. C: Structural connectivity associated with axial improvement under DBS. Stimulation of the cerebellothalamic (dDRT) and corticospinal tracts (CST), as well as subthalamic afferents from the globus pallidus externus (indirect) positively correlated with outcome, while the opposite was observed for the hyperdirect pathway from the primary motor cortex (HDP-M1). D: Voxel-wise correlation map of the electric field magnitude with stimulation outcome. The sweet-spot, thresholded for significance, is primarily located in the ventral oral posterior nucleus of thalamus impinging on the internal capsule, while the sour-spot is identified at the border of the motor and associative STN. E: Clinical scores and their association with the fiber filtering and sweet-spot mapping models. To indicate robustness of the association, the leave-one-out cross-validation is reported. For 10-fold see panel G. The models were also subjected to permutation tests: R = 0.61, p < 0.01 and R = 0.55, p > 0.05 for the sweet-spot and fiber filtering models, respectively. F: Peaks of the functional correlation map for cervical dystonia primarily reside in the cingulo-opercular network, delineated in gray contours. G: Boxplots of 10-shuffles of 10-fold cross-validation.*

### Supplementary Figure 3: Correlation of improvements across items

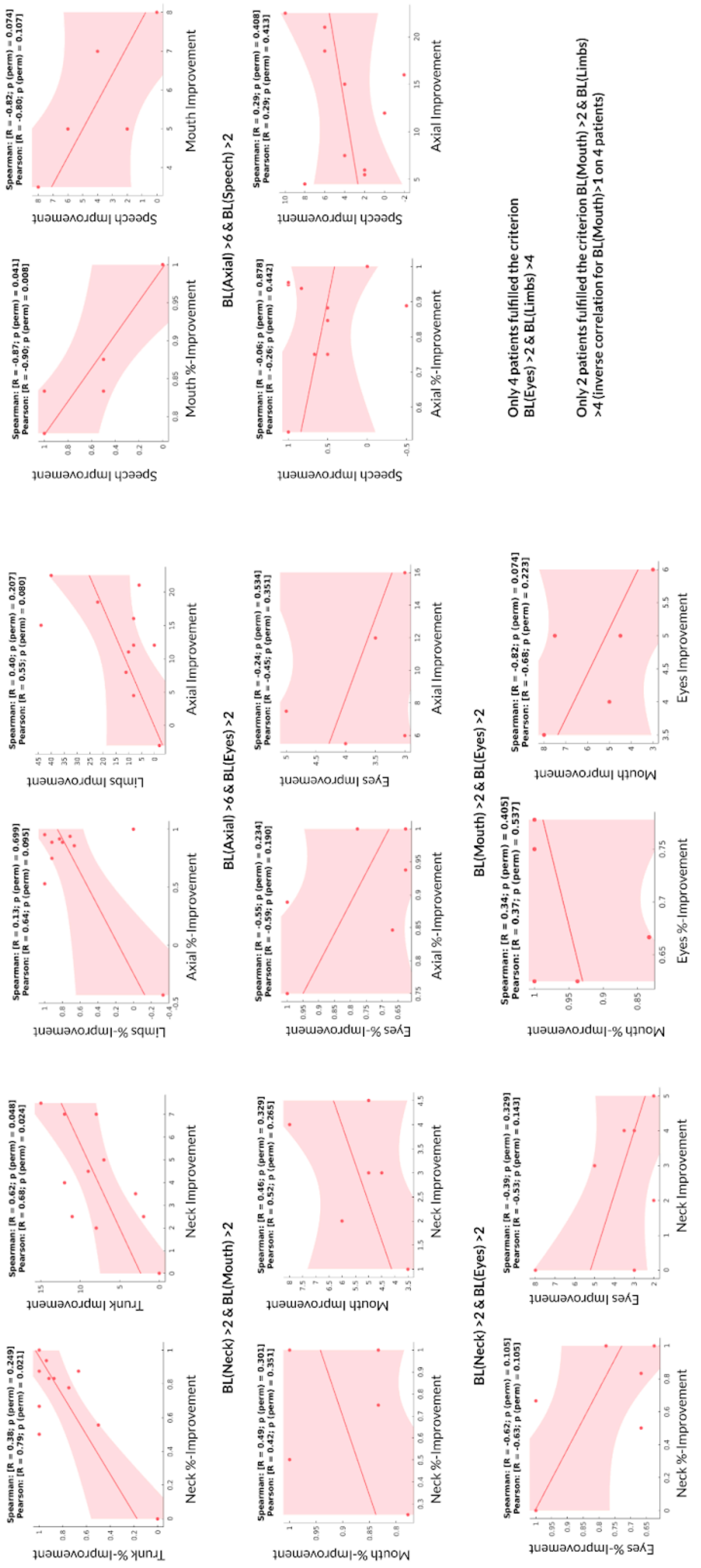

### Supplementary Table 4: Patient demographics

| | N (female) | Age at Surgery, years | Disease Duration, years | Follow-up, years | %-Improvement* |
|---|---|---|---|---|---|
| All patients | 58(29) | 40.2 ± 20.1 | 6.6 ± 7.6 | 1.3 ± 1.2 | 67.7 ± 28.4 |
| Cervical group | 30(12) | 41.6 ± 20.1 | 6.9 ± 7.3 | 1.0 ± 0.8 | 64.4 ± 34.9 |
| Appendicular group | 27(10) | 29.6 ± 19.3 | 8.4 ± 9.9 | 1.3 ± 1.1 | 73.7 ± 32.1 |
| Blepharospasm group | 21(12) | 47.8 ± 18.9 | 5.1 ± 5.4 | 1.4 ± 1.4 | 82.3 ± 13.1 |

*BFMDRS percent improvement is reported for all patients, for groups - percent improvement for the corresponding BFMDRS items.

### Supplementary Figure 5: Cross-correlation of the disease duration / age at surgery with outcome for appendicular, cervical and blepharospasm improvement

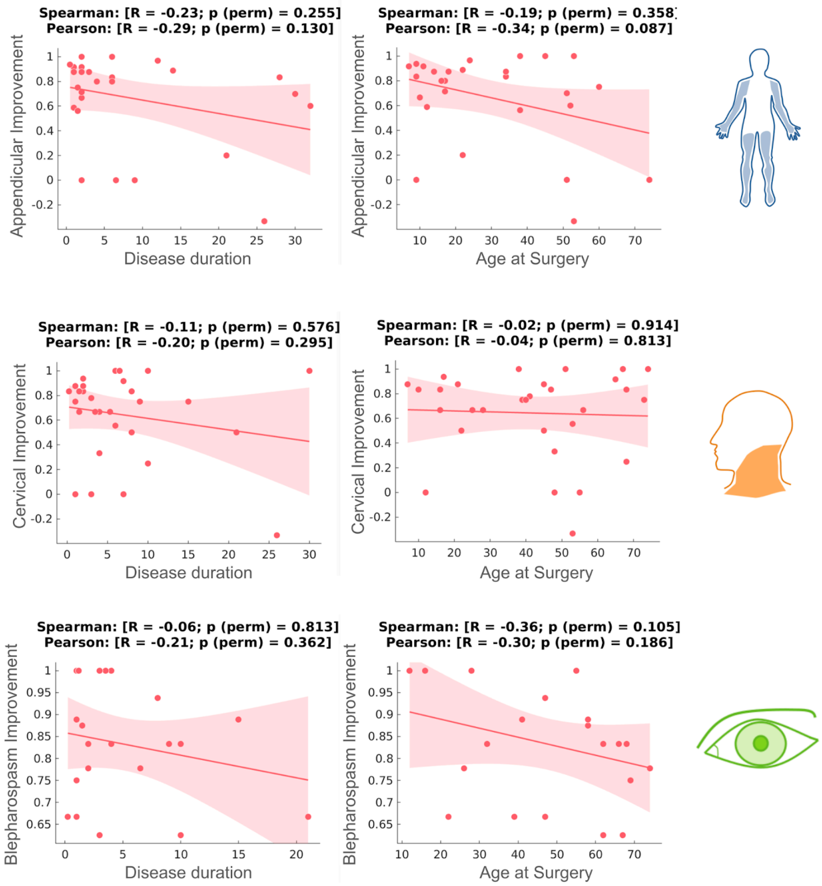

### Supplementary Figure 6: Cross-correlation of appendicular and blepharospasm models

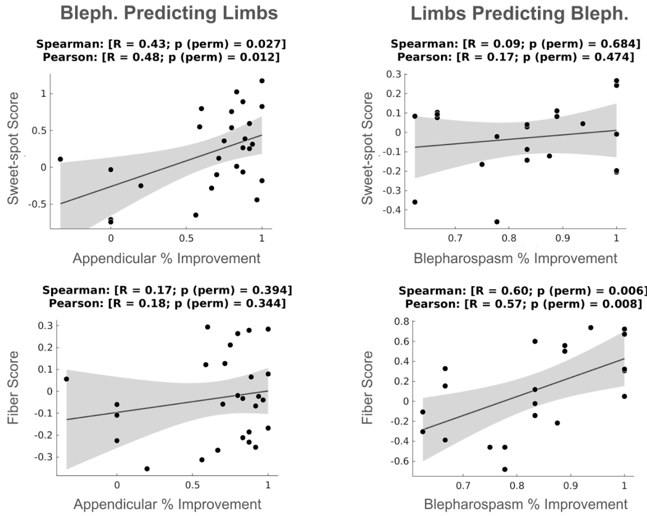
